# Spatial Distribution of Missense Variants within Complement Proteins Associates with Age Related Macular Degeneration

**DOI:** 10.1101/2023.08.28.23294686

**Authors:** Michelle Grunin, Sarah de Jong, Ellen L Palmer, Bowen Jin, David Rinker, Christopher Moth, Anthony Capra, Jonathan L. Haines, William S Bush, Anneke I. den Hollander, IAMDGC

**Author notes:** indicates equal contribution (Michelle Grunin, Sarah de Jong, Ellen L Palmer, Jonathan L. Haines, William S Bush, Anneke I. den Hollander). indicates corporate author (IAMDGC).

## Abstract

**Purpose:** Genetic variants in complement genes are associated with age-related macular degeneration (AMD). However, many rare variants have been identified in these genes, but have an unknown significance, and their impact on protein function and structure is still unknown. We set out to address this issue by evaluating the spatial placement and impact on protein structureof these variants by developing an analytical pipeline and applying it to the International AMD Genomics Consortium (IAMDGC) dataset (16,144 AMD cases, 17,832 controls).

**Methods:** The IAMDGC dataset was imputed using the Haplotype Reference Consortium (HRC), leading to an improvement of over 30% more imputed variants, over the original 1000 Genomes imputation. Variants were extracted for the *CFH*, *CFI*, *CFB*, *C9*, and *C3* genes, and filtered for missense variants in solved protein structures. We evaluated these variants as to their placement in the three-dimensional structure of the protein (i.e. spatial proximity in the protein), as well as AMD association. We applied several pipelines to a) calculate spatial proximity to known AMD variants versus gnomAD variants, b) assess a variant’s likelihood of causing protein destabilization via calculation of predicted free energy change (ddG) using Rosetta, and c) whole gene-based testing to test for statistical associations. Gene-based testing using seqMeta was performed using a) all variants b) variants near known AMD variants or c) with a ddG >|2|. Further, we applied a structural kernel adaptation of SKAT testing (POKEMON) to confirm the association of spatial distributions of missense variants to AMD. Finally, we used logistic regression on known AMD variants in *CFI* to identify variants leading to >50% reduction in protein expression from known AMD patient carriers of CFI variants compared to wild type (as determined by *in vitro* experiments) to determine the pipeline’s robustness in identifying AMD-relevant variants. These results were compared to functional impact scores, ie CADD values > 10, which indicate if a variant may have a large functional impact genomewide, to determine if our metrics have better discriminative power than existing variant assessment methods. Once our pipeline had been validated, we then performed *a priori* selection of variants using this pipeline methodology, and tested AMD patient cell lines that carried those selected variants from the EUGENDA cohort (n=34). We investigated complement pathway protein expression *in vitro*, looking at multiple components of the complement factor pathway in patient carriers of bioinformatically identified variants.

**Results:** Multiple variants were found with a ddG>|2| in each complement gene investigated. Gene-based tests using known and novel missense variants identified significant associations of the *C3*, *C9*, *CFB*, and *CFH* genes with AMD risk after controlling for age and sex (P=3.22×10^−5^;7.58×10^−6^;2.1×10^−3^;1.2×10^−31^). ddG filtering and SKAT-O tests indicate that missense variants that are predicted to destabilize the protein, in both CFI and CFH, are associated with AMD (P=CFH:0.05, CFI:0.01, threshold of 0.05 significance). Our structural kernel approach identified spatial associations for AMD risk within the protein structures for C3, C9, CFB, CFH, and CFI at a nominal p-value of 0.05. Both ddG and CADD scores were predictive of reduced CFI protein expression, with ROC curve analyses indicating ddG is a better predictor (AUCs of 0.76 and 0.69, respectively). *A priori in vitro* analysis of variants in all complement factor genes indicated that several variants identified via bioinformatics programs PathProx/POKEMON in our pipeline via *in vitro* experiments caused significant change in complement protein expression (P=0.04) in actual patient carriers of those variants, via ELISA testing of proteins in the complement factor pathway, and were previously unknown to contribute to AMD pathogenesis.

**Conclusion:** We demonstrate for the first time that missense variants in complement genes cluster together spatially and are associated with AMD case/control status. Using this method, we can identify *CFI* and *CFH* variants of previously unknown significance that are predicted to destabilize the proteins. These variants, both in and outside spatial clusters, can predict *in-vitro* tested *CFI* protein expression changes, and we hypothesize the same is true for *CFH*. *A priori* identification of variants that impact gene expression allow for classification for previously classified as VUS. Further investigation is needed to validate the models for additional variants and to be applied to all AMD-associated genes.

## Introduction

Changes to protein structures are a common cause of Mendelian disease, and are often found to influence risk of complex diseases, including age-related macular degeneration (AMD)^1^. Traditional computational methodologies for scoring protein structure impact are done via modeling known protein structures and identifying their predicted impact based on known functional variants (e.g. Polyphen/Sift^2^). These predictions are most accurate when a variant in question is located in a high impact area of the protein and involve a dramatic amino acid substitution or deletion, such as variants located in an alpha helix or beta sheet^3–5^. However, the impact of more minor changes, such as missense mutations in unstructured regions or with a less dramatic amino acid substitution, on protein function is less clear and has proven difficult to evaluate. One approach to address this issue is to calculate free energy change, but these are computationally intensive^6^, and are not generally performed on a large scale, though some thermodynamic changes can be found in the database ProTherm^7^. In addition, only solved protein structures from the Protein Data Bank (PDB)^8^ can be utilized for computational evaluation of both minor and major changes. As of 2020, 14,028 solved structures for proteins can be found in the PDB, although non experimentally and predicted structures can be created with modeling methods like Swiss-Model utilizing the Expasy webserver^9^. In 2021, AlphaFold was released to the public, allowing for most structures to be solved with great accuracy due to the machine learning algorithm, leading to the fact that most structures are available for use to evaluate changes (Jumper, 2021). In addition, the identification of a method that can reliably identify those factors that impact stability that then impact protein function, especially in disease, has not been determined, despite the solved protein structures available in the PDB and through AlphaFold.

The complement factor pathway is well known to play a role in AMD pathogenesis^10^. Variation in multiple genes in the complement factor pathway (e.g. CFH, C9, C3, CFB) are known risk loci, as well as possible drug therapy targets. The International Age-related Macular Degeneration Genomics Consortium (IAMDGC) identified and confirmed variants in the complement factor genes that influence AMD^10^, and further analyses have identified rare variants in the complement factor genes that influence protein expression and function^11–20^ However, it is unknown if all variants in those genes contribute to pathogenesis of AMD, or how those variants impact protein expression and function. Classification of variants of unknown significance (VUS) has clinical impact, and it is unclear if rare variants, which may not be powered for detection in a GWAS, influence AMD pathogenesis, or if there are criteria that we can use to identify only those variants that influence protein expression or structure.

In addition, many of variants in the other 29 previously AMD-associated loci have not well evaluated as to protein expression in a large scale. Further, it is unknown if the functional mechanisms of the rare variants in the complement genes, influence AMD pathogenes in the same way as the well-known common variants (e.g. Y402H in CFH). These variants may not influence protein structure in obvious ways, but rather cause minor distortions to binding sites, pockets of influence (like in gating areas or ion channels), and stability in more subtle ways. Identifying likely functional variants from those of unknown significance in both the complement genes and the other AMD genes would allow for further identification of important areas of the protein for drug targets and therapy. Leveraging protein spatial proximity to known AMD variants, like the 52 already known variants^10^, would allow classification of variants as detrimental or neutral.

Once variants affecting protein structure can be identified computationally, the identification of variants that influence protein expression in actual bench experiments can be performed. Variants that influence CFI expression have been detailed previously^15^, and we can validate our computational identification by looking at protein expression in previously tested variants.. We aim to perform the “holy grail” of variant testing, to winnow down large scale listings to variants than can be computationally predicted and experimentally validated. Using this pipeline, we aimed to see if we could replicate identification of variants where we already knew their impact on protein expression, and then move one step further to predict which novel variants, not previously associated with AMD pathogenesis, would have a similar effect on protein expression in the complement factor pathway. This would be a proof-of-concept whether we can *a priori* identify variants computationally as likely damaging that affect protein expression experimentally *in vitro*.

We hypothesize that the spatial proximity within a protein between variants of known significance and rare variants of unknown significance can help predict the functionality of these uncharacterized variants.We tested these variants’ impact on protein structure and function using the genotyped data of the complement factor genes in AMD. We utilize PathProx^21,22^ to evaluate the spatial proximity between known disease-causing variants and variants observed in the IAMDGC and POKEMON^23^ to evaluate clustering of variants found to be more prevalent in cases alone, or controls alone. Finally, computationally modeling of the change in free energy needed to fold the protein in the presence of these missense variants using Rosetta provides a comprehensive picture of the likely role of these variants in AMD risk.

## Methods

### Genetic data on AMD

The IAMDGC dataset (original: AMD=16,144, control=17,832) has 250K tagging and 250K rare/common variants on a custom chip developed for the IAMDGC (Affymetrix, detailed in Fritsche et al 2016). The starting point for pre-imputation was 569,645 variants genome wide genotyped on the custom chip. These were filtered according to previous methodology for SNPs that were problematic due to whole genome amplified samples, sex-specific associations, or variants that did not genotype well according to QC parameters described in Fritsche et al (2016). Briefly, only individuals with a known phenotype (geographic atrophy (GA), neovascular AMD (nvAMD) and dry/atrophic AMD (aAMD), and European descent based on the previous population stratification analysis^10^ were used. Analysis was performed with Plink 1.9 and XWAS 3.0, along with R and Bioconductor. Of the IAMDGC, 18,865 female and 10,404 male samples were used (12,087 control/14,273 AMD). All data collection was approved by the institutional review board or ethics committee of all participating institutions.

The Haplotype Reference Consortium panel (HRC) 1.1 dataset was used for the imputation reference panel^24^. It contained 64,976 haplotypes including chromosome X. The Michigan Imputation Server was utilized, and ShapeIT^25^ was used for pre-phasing(HG19 data). Imputation was performed using minimac3^26^.

The final variant count was 275,759 before imputation after filtering and QC. The QC pre-imputation included a identity by descent calculation (IBD), removal of the heterozygote haploid genotypes on X, Hardy-Weinberg equilibrium (HWE) calculated separately for males and females and the variants removed, missing and frequency testing was performed according to standard and variants removed, along with removal of samples with missing phenotypes. 3 SNPs failed the sex frequency testing at a P=7.56×10^-6, and were removed.

All samples were then imputed using HRC 1.1. Post-imputation processing sensitive to gender occurred in similar steps to pre-imputation processing including HWE, missingness, frequency and sex-specific details, along with the QUAL threshold. There were still some SNPs (n=1273) with a different frequency amongst sexes in controls with P<5.9×10^−8^ which were removed. The improvement in imputation was 3-fold the number of variants successfully imputed compared to the original IAMDGC data.

Data for CFH, CFI, C3, C9, and CFB were pulled after HRC 1.1 imputation. 500 base pairs before and after gene start/end were pulled to include the maximum amount of variants available. Hg19 was used for reference. All sample phenotypes were used, and then filtered for advanced AMD, which includes both neovascular AMD and geographic atrophy, the same definition for advanced AMD as in Fritsche et al.^10^. Each gene contained a different number of variants pulled from the IAMDGC data within the boundaries listed below.

### Risk and ‘neutral’ variant identification

A literature review for all AMD associated variants in the complement factor complex was completed on [30.11.2020]. Missense variants were retained for algorithm usage while nonsense, splice site, ncRNA, near gene, or UTR variants were excluded. We utilized the genome aggregation database v2.1.1 (gnomAD)^28^ to create lists of likely neutral variants. GnomAD provides summary statistics from 125,748 exome sequences and 15,708 whole-genome sequences of unrelated individuals who have served as controls in various disease and population genetic studies.

Pipeline for Spatial Associations:

We developed a pipeline to identify spatial associations from the lists of variants with unknown pathological classsication utilizing both POKEMON and PathProx. Each step included both

### Spatial assessments

Assessments were performed according to the same methodology as Jin, et al (2022). The methods in brief: Protein structures that were both experimentally solved and those that could not be experimentally solved were predicted, and those structures were taken from the structures in the Protein Databank (PDB) [29], and from both Swiss Model and ModBase. Each gene underwent cross-referencing to the structures, via UniProt ID Mapping [30], and the Swiss Models [8] were cross referenced with UniProt identities as well. ModBase [31] were downloaded from ftp://salilab.org/databases/modbase and Ensembl was used to cross-reference the proteins and their transcripts. Analysis was performed on single-chain monomers. The PDB chains were aligned to canonical UniProt transcripts with SIFTS [32], and aligned to non-canonical transcripts with the SIFTS REST API. If multiple structures existed, the structure curation was guided heuristically to maximize diversity, coverage around variant positions of interest, resolution of models, and the sequence identity.

### Spatial assessments

In the spatial comparisons of variants without prior pathogenic knowledge to AMD to characterized variants (PathProx)^21,22^, pathogenic variants were taken from ClinVar and neutral variants from ExAC 2.1^29^, and variants were assigned the 3D location of the centroid of the modeled amino acid side chains. Each variant position was weighted with normalized BLOSUM^30^ 100 scores, ranging of 0 to 1. Variants with a PathProx score > 0 are on average closer in 3D to observed pathogenic than neutral variants, while variants with a PathProx score < 0 are on average more proximal to neutral variants.

Computational evaluation on the effects of a variant on the free energy of folding of protein was carried out using ΔΔG Monomer from the Rosetta biomolecular modeling suite^31^. For X-ray structures of 2.5 Å resolution or better the “Row 16”^19^ protocol from Kellogg *et al*.^32^ was used; “Row 3” was used otherwise. ΔΔG Monomer calculations were only performed for Swiss and ModBase models with at least 40% sequence identity to their PDB template structures. For the purposes of our work, a |ΔΔG| > 2 is considered extreme enough to potentially impact protein stability.

### Gene-based testing

Previous studies have shown that removing neutral variants from gene-based testing ^21^ may improve the potential for integration of genomics pipelines into eventual clinical assessment [To maximize the benefit of gene-based testing, we assessed each set of variants (PathProx and ddG) using seqMeta, an algorithm that identifies the optimal rho between burden and SKAT testing^33^. (R package version 1.6.7. 2017 https://CRAN.R-project.org/package=seqMeta.). In summary, variants with a PathProx score > 0 (demonstrating a closer proximity to known risk variants over neutral variants) or a ddG > 2 (predicted to destabilize the protein structure) were retained for gene-based testing. All variants that failed to meet one or more criteria were presumed putatively neutral and excluded from all downstream analyses.

We also employed an orthogonal approach as per Jin et al^34^, to determine if there was a spatial relationship between variants and case-status. This method uses a structural-kernel-based variance component test to incorporate spatial proximity between rare variants when calculating the gene-based statistic. In short, if two individuals have variants that are proximal to each other within the protein structure, they are considered genetically more similar than two people who have variants at opposite ends of the 3D protein structure. This structural-kernel is then used for subsequent gene-based testing to determine if there are differences in the spatial patterns of variants between cases vs. controls.

Finally, to give additional context to our findings we calculated the seqMeta statistics for all missense variants in the gene. While not directly comparable to the results from the above testing due to changes in sample size between assessments, these statistics provide additional contextual information for interpreting the PathProx and ddG results. For a direct assessment of potential enrichment of, we calculated the case carrier percentage for each test: all missense, PathProx > 0, or ddG > 2. We report all observed p-values, and consider a nominal p < 0.05 indicative of observed spatial clustering.

### Prediction of detrimental CFI secretion changes

To determine if these spatial assessments provide information that can be used to both understand and predict underlying biology, we chose to further investigate the *CFI* gene, which has a large body of literature about the impact of specific variants on gene expression. Using our previously published data on the effect of mutant *CFI* on Factor I secretion levels^35^ we retained all missense variants that were found in CFI after classification of those variants as missense. PathProx and ddG scores were calculated for these variants. These data were then randomly split into 80/20 training/testing sets, with 4-fold cross-validation done on the 80% training set and a final model performance assessment completed in the 20% never used in training.

To create a binary classifier related to known biological function that could be readily deployed clinically, we declared that a reduction in CFI expression to <50% that of the canonical transcript was detrimental and chose this as our cutoff of ‘functional’ vs. ‘not functional’. We assessed models using both the numerical outputs of PathProx, ddG, and CADD (for comparison to existing variant scoring methods) and binary classifiers (PathProx > 0, ddG > 2, and CADD > 10 representing detrimental variants). We ran unadjusted models and models adjusting for age, sex, age+sex, and all combinations of the three variant function predictors.

### Validation in plasma and serum samples of rare variant carriers

Carriers of likely damaging variants were selected from the European Genetic Database (EUGENDA) for validating the predicted effect in plasma and serum samples. In total 35 rare variant carriers were included (Supp. Table 1), and the concentrations of complement components were determined in EDTA plasma with enzyme-linked immunosorbent assay (ELISA). We investigated Factor I and the complement component C3bBbP which is a protein found in the end of the alternative complement pathway and is an indication of complement activation. In two *CFI* variant carriersFI and C3bBbP were measured, in two *C3* variant carriers C3bBbP was measured, in nine *CFB* variant carriers FB and C3bBbP were measured, and in 21 *C9* variant carriers terminal C5b-9 complex (TCC) was measured in plasma, and the lytic activity was determined in serum with the hemolytic assay. Measurements were performed in the routine diagnostics setting in the Department of Laboratory Medicine of the Radboudumc. The ELISA procedures are summarized in Supp. Table 2. The hemolytic assay was performed according to the following: Rabbit erythrocytes (Envigo, Netherlands) were washed and diluted with magnesium-ethylene glycol tetraacetic acid (Mg-EGTA) buffer (2.03 mM veronal buffer, pH 7.4, 10 mM EGTA, 7 mM MgCl2, 0.083% gelatin, 115 mM D-glucose, and 60 mM NaCl) and diluted in a way, that the absorbance of fully lysed erythrocytes was between 0.8-1.2 at 405 nm. Serum samples were diluted in Mg-EGTA buffer, and 20 ul of diluted serum were mixed with 10 ul erythrocytes and incubated at 37C and 600rpm for 30 min. The reaction was stopped with 150ul EDTA-GVB, and the samples centrifuged for 2 min at 1000g to remove intact erythrocytes. The amount of lysis was determined by measuring the absorbance in supernatants at 405 nm. For FI, FB and sTCC and the hemolytic activity the default diagnostic reference range was used, while for C3bBbP a cohort specific reference range was determined based on de Jong et al. 2022. ^20^

**Table 1:**
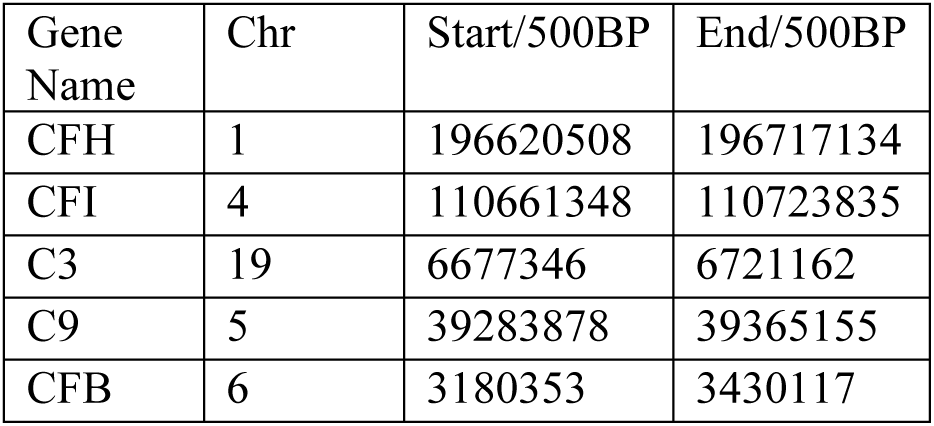
Different amount of missense variants pulled per complement gene (hg37)

**Table 2:** Calculation of phenotype with and without inclusion of covariates via POKEMON on all complement genes tested.

| GENE | PDB | PHENOTYPE | P-VALUE |  |  |
| --- | --- | --- | --- | --- | --- |
|  |  |  | No covariates | Covariate:SEX | Covariate:SEX and AGE |
| GeneC3 | 2a73 | CNV_only vs No_AMD | 0.18 | 0.18 | 0.17 |
| GeneC9 | 6h04 | CNV_only vs No_AMD | 9.7E-03 | 0.015 | 0.09 |
| GeneCFH | 1haq | CNV_only vs No_AMD | 1.1E-06 | 1.4E-06 | 2.1E-05 |
| GeneCFI | 2xrc | CNV_only vs No_AMD | 0.13 | 0.15 | 0.25 |
|  |  |  | No covariates | Covariate:SEX | Covariate:SEX and AGE |
| GeneC3 | 2a73 | (CNV_only, Mixed_GA/CNV, and GA_only) vs No_AMD | 0.10 | 0.10 | 0.09 |
| GeneC9 | 6h04 | (CNV_only, Mixed_GA/CNV, and GA_only) vs No_AMD | 4.9E-03 | 6.6E-03 | 0.04 |
| GeneCFH | 1haq | (CNV_only, Mixed_GA/CNV, and GA_only) vs No_AMD | 2.4E-09 | 2.4E-09 | 2.7E-09 |
| GeneCFI | 2xrc | (CNV_only, Mixed_GA/CNV, and GA_only) vs No_AMD | 1.06E-02 | 1.11E-02 | 8.46E-03 |

## Results

### AMD genetics data sources and imputation with HRC

The full set of the IAMDGC data was imputed with the Haplotype Reference Consortium (HRC v1.1). SHAPEIT was used for phasing and minimac3 for imputation. After imputation with the HRC more than 120 variants were pulled for each gene in the complement factor pathway (CFH, CFI, C3, C9, CFB). (Table 1). No variants were excluded from any sample that was tested, and variants were all genotyped or well imputed (R^2^ > 0.8).

### Spatial Assessments

In total, 323 out of 659 (49%) variants were located in protein structures of sufficient quality to assess ddG as per the methods originally, utilizing both Ensembl and UniProt transcripts, and utilizing the three structure databases detailed in Methods. A total of 184 (27.9%) had an extreme ddG (>=2) suggestive of unfavorable protein folding; 64 of these variants were in *C3*, 10 in *C9*, 63 in *CFH*, and 47 in *CFI*. These variants identified as unfavorable for protein folding via this methodology were classified as PathProx positive (PP+) For the same 323 variants in analyzable protein structures, four variants in C3 were PP+C9 also contained four PP+ variants, 53 were PP+ in CFI, and 13 in CFH. There were no variants in CFB classified as PP+ and this gene was removed from subsequent analyses in PathProx but not in POKEMON.Due to the nature of POKEMON, all variants available in structures were included in that analysis to identify potential clusters associated with case-status.

### Gene-based testings

PathProx SKAT-O results on those variants that were PP+ indicate that novel observed variants in CFH, CFI, and C9 that are proximal to known AMD variants (taken from the Fritsche et al, 2016 paper^10^) are associated with AMD risk (P=CFH=0 (threshold<1×10^-15), CFI=0.01, C9=0.03). PathProx testing of C3 did not identify any variants in close proximity to known pathological variants(C3=0.83)). Since destabilization of CFH and CFI is thought to be causally associated with AMD and our statistical test found the unknown variants were associated with AMD risk, we further assessed these two genes to see if variants predicted to impact protein folding (using ddG) were similarly associated with the AMD phenotype.

SKAT testing utilizing only variants with a notable impact on ddG found that variants predicted to destabilize CFI and CFH are associated with AMD (CFH P=0.05, CFH P=0.01). The other complement genes did not show statistical significance with free-energy SKAT testing alone (C3 P=0.33, C9 P=0.09)In addition, grouping of ddG variants into a SKAT test implicitly assumes that a high ddG variant is damaging whether found in a highly structured or disordered protein domain, potentially weakening a true association.

The POKEMON method, an orthogonal test from PathProx that does not rely on external data sources, was also evaluated in these five complement genes. All complement genes have clusters of variants in the protein structure that were more associated with cases or controls than expected by chance alone (Kernel+frequency testing POKEMON P: C3=0.04, C9=0.01, CFB=2.84×10^−14^, CFH=0 (threshold<1×10^-15), CFI=0.02). When common variants were used, we performed inverse scoring on the variants according to the count of people with said variants, so we can use frequency to lower the effect that the common variants would have according to the frequency, not just spatial distribution. Case-associated clusters were found at the terminal end of the CFH (Figure 1) while control-associated variants were more commonly found through the center of CFH. Similarly, for C9, 69 variants more commonly found in controls were located in the external ring of the protein, while in the internal passageway, the identified variants were more commonly found in cases (Figures 1–2).

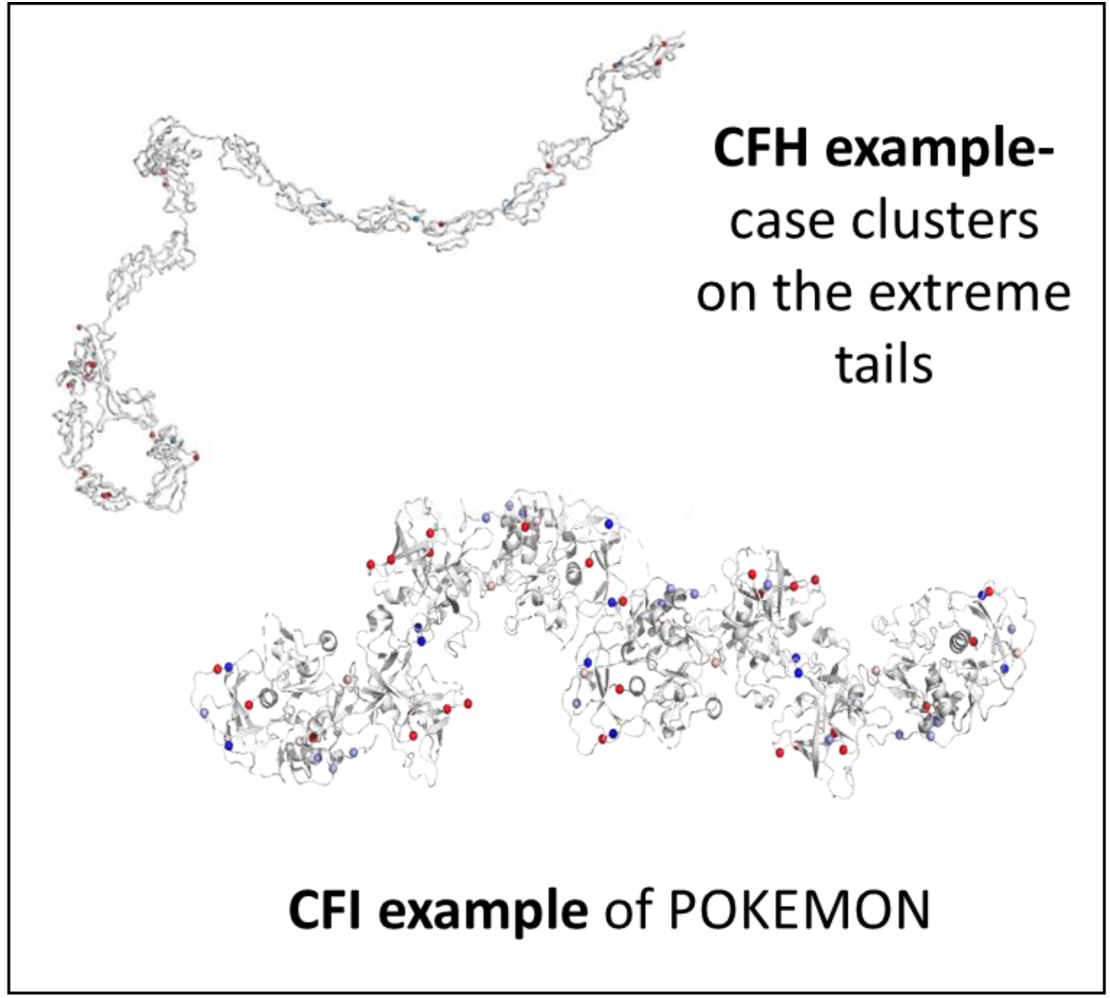
Figure 1.

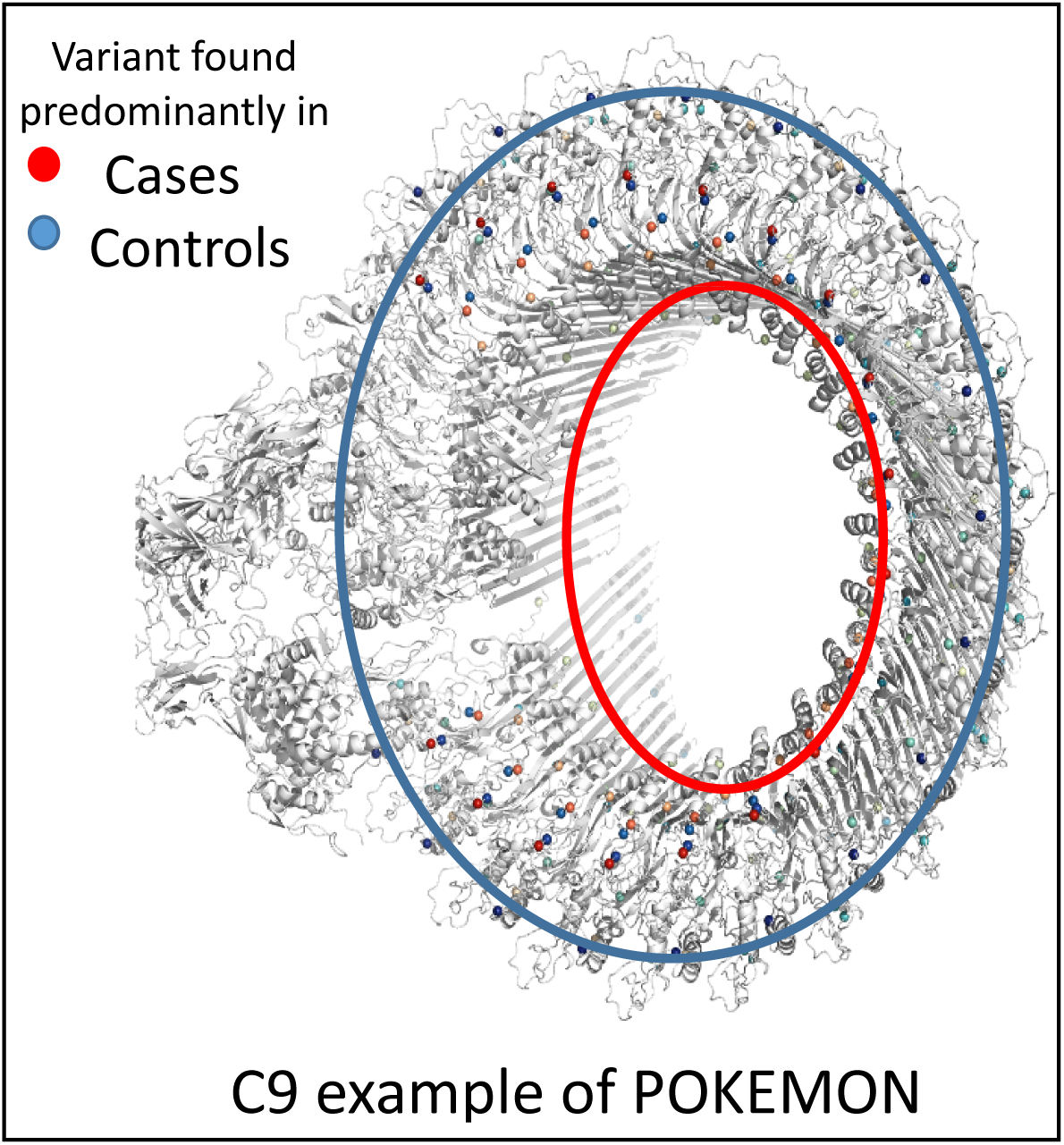
Figure 2.

After removing known associated variants, POKEMON was applied to the subphenotype of choroidal neovascularization (CNV) cases only. The association with CFH was significant after application of both sex and age as covariates (Table 2 Section 1). When applied to all advanced AMD cases, after including sex and age, C9, CFH, and CFI still indicated significance, showing that although the covariates of sex and age still have an effect, they do not temper the clusters of case/control variants found in the protein (Table 2 Section 2).

### Prediction of detrimental CFI expression changes

We assessed if the functional predictions associated with previously bench validated functional changes in CFI expression, as detailed in de Jong et al 2020^15^. We already knew that more than 50% of these variants affect protein secretion, but for confirmation we performed association analysis with variants that were identified via our spatial analysis.

ROC curve analysis indicated that ddG was a predictor of CFI expression change, with an AUC of 0.77 vs the CADD score model (Figure 3 A indicating CADD score effectiveness compared with our current ROC curve analysis in Figure 3 B) with an AUC of 0.59 (a known benchmark of predicted functionality). Further, this analysis had a sensitivity of 0.78 and a specificity of 0.54 indicating it has clinical value. Further details can be found in Figures 3–4 and Supplementary Figure 1A-D.

**Figure 3:**
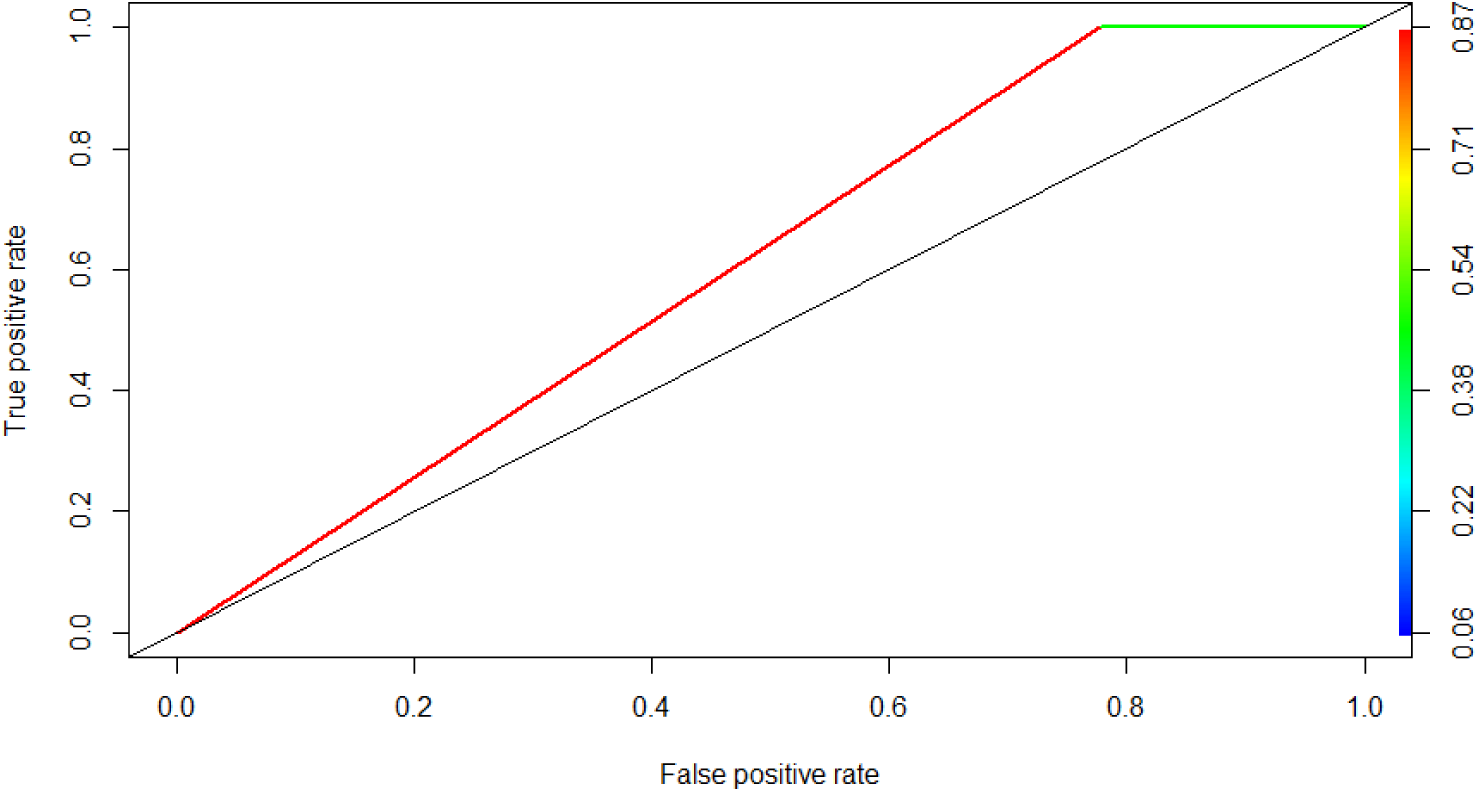
CADD scores (binary)

**Figure 4:**
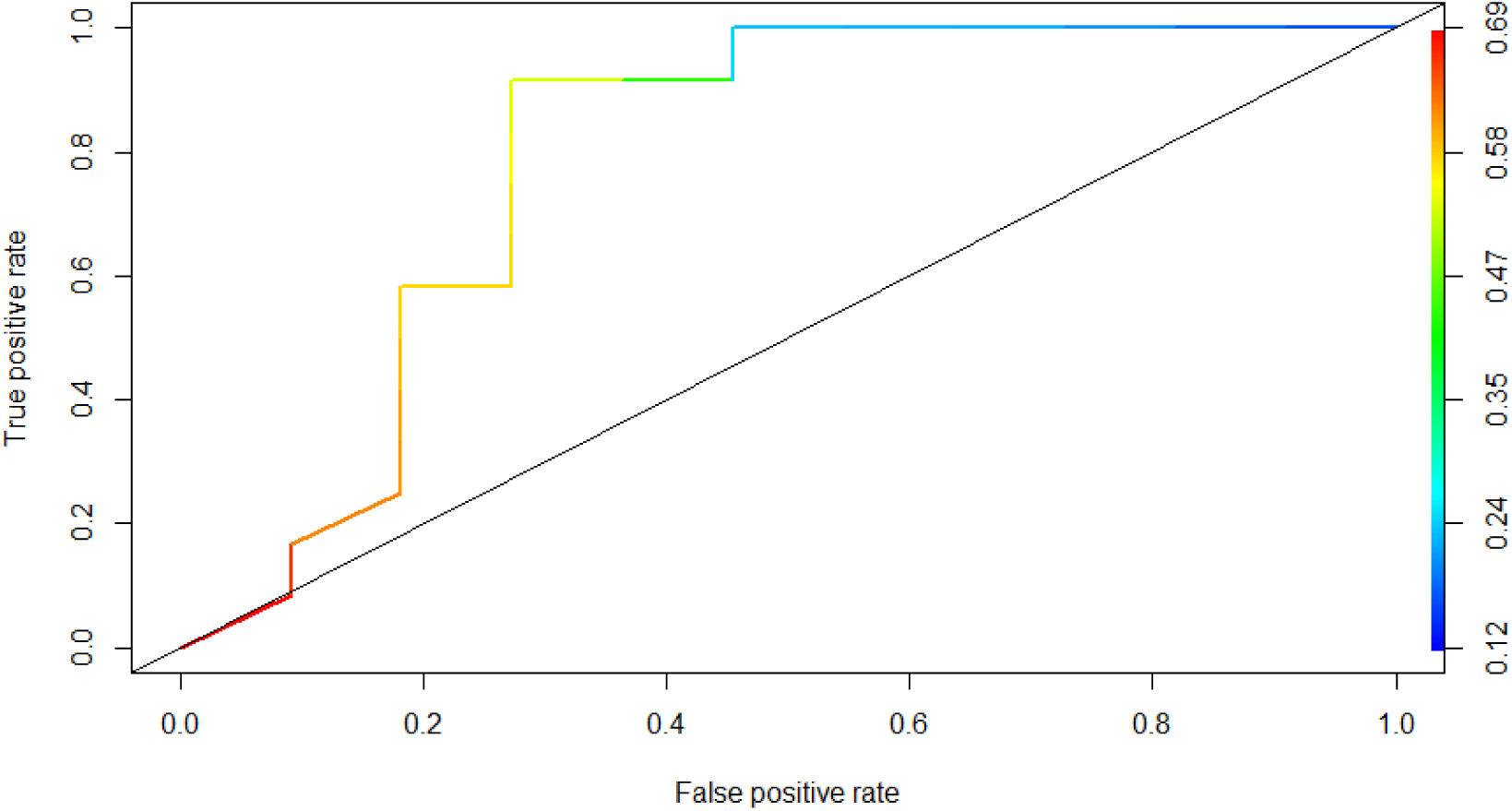
ddG binary and CADD (continuous)

### Validation in plasma and serum samples of rare variant carriers

After the indication that we could predict FI secretion, we set out *a priori* to examine protein levels in plasma of rare variant carriers selected from the EUGENDA database. We performed *a priori* hypothesis testing, predicting that these variants would affect complement gene expression at a rate greater than random chance.

In total we identified 34 carriers of variants predicted to affect the protein stability that could be included for functional testing. Of these 12 carriers presented with no AMD, 10 presented with early/ intermediate AMD, 11 presented with advanced AMD, and one individual could not be graded. The average age was 74.7±1.24. The measurements performed as outcome were taken at different points of the complement factor pathway: C3bBbP (which was also utilized as a threshold for significant difference, which was indicated at < 19 CAU/ml), CFI, CFB, and TCC (which is the end marker for the complement gene pathways, and it does not always signal completely in clinical experiments. (Supplemental Table 3).

Results indicate that despite the small sample size (n=34), there is a significant difference in protein expression *in vitro*, in the complement pathway amongst carriers of the variants identified in our spatial variant methodology. For example, out of 14 variant carriers for CFB G252S, 9 carried above normal expression of C3bBbP (P=0.04, Student’s T-Test). Of those, 2 of those 9 also carried abnormal CFI expression, and 2 contained abnormal CFB expression (Table 3, Supplementary Table 4).

**Table 3:** Indication of patients with their age, phenotype and significantly impacted complement pathway expression results.

| Patient ID | Phenotype | Age | C3bBbP CAU/ml | CFI (ug/ml) | CFB (ug/ml) |
| --- | --- | --- | --- | --- | --- |
| Patient 5 | Advanced AMD: ne | 72.21 | 21.1 | 19 |  |
| Patient 4 | Advanced AMD: ne | 72.73 | 23.8 |  | 173 |
| Patient 2 | Advanced AMD: ne | 74.1 | 19.8 |  | 292 |
| Patient 6 | Advanced AMD: ne | 77.21 | 21.6 | 20 |  |
| Patient 1 | Advanced AMD: ne | 81.63 | 27.4 |  |  |
| Patient 3 | Advanced AMD: ne | 85.26 | 196 |  | 186 |
| Patient 7 | cannot grade | 72.25 | 16.4 |  | 180 |
| Patient 9 | early AMD | 80.63 | 13.6 |  | 213 |
| Patient 8 | early AMD | 81.12 | 17.6 |  |  |
| Patient 10 | intermediate AMD | 75.35 | 22.9 |  | 171 |
| Patient 11 | no AMD | 74.64 | 16.9 |  | 184 |
| Patient 13 | no AMD | 75.2 | 13.4 |  | 170 |
| Patient 14 | no AMD | 77.93 | 24.4 | 36 |  |
| Patient 12 | no AMD | 78.43 | 147 |  | 181 |

## Discussion

We have shown, using complementary novel statistical methodologies, that variants of unknown significance (VUS) can be identified that impact protein conformation and protein expression. This computational testing can identify, from a list of observed variants, which ones are predicted via spatial modeling to have an effect, allowing for more efficient prioritization of bench testing novel observed variants.

This computational pipeline was tested on the complement genes most well known in AMD, and our results were compared to previously published values of gene expression in CFI. After confirming that we could reliably identify variants with an observed impact on the expression of CFI, we predicted which novel observed variants not previously associated with AMD would have a similar impact on gene expression, identifying novel variants meeting this threshold. When we combine the free energy results with spatial relationships, we show that these variants of previously unknown significance (VUS), have damaging effects, and are associated with AMD pathogenesis. We have demonstrated that variants, computationally identified as likely damaging, affect protein expression *in vitro.* This novel methodology can be applied to any disease.

In CFH we observe a POKEMON cluster in CCP1, further significant associations were identified in CCP3, 4 and 20. Rare coding variants in CFH have been reported to affect binding capacities or co-factor function rather than affecting CFH secretion levels, but we have recently identified significantly lower CFH plasma levels in rare variant carriers compared to controls^20^ In line with this, we observe here a significant association of destabilizing CFH variants in AMD patients, and 63 out of 114 (55%) variants are predicted to destabilize CFH with a ddg score larger than 2. It should be noted, however, that the resolution of the CFH protein structure is limited by the flexibility of the protein, potentially affecting our analysis. Also, the two coding variants in CFH showing the highest association with AMD, p.Tyr402His and p.Arg1210Cys, show altered binding capacities to extracellular matrix components or albumin binding respectively. These effects are not detected in our analyses and present a limitation of the model in context of interpreting variants in CFH. A limitation of our study is that these results are only as good as the solved protein structure, indicating that there are some resolution limitations, however many proteins have been successfully mapped in in crystal structure, indicating that solved protein structures are accurate.

Rare coding variants in CFI affect FI secretion levels ^12,15,17^, with more than half of the studied variants affecting FI secretion. Here we observe that 47 out of 85 (55%) CFI variants are predicted to destabilize the protein with a ddG score above 2, and we show an increased AUC of 0.77 compared to the commonly used CADD score (AUC 0.59). Pokemon analysis shows distribution of destabilizing clusters associated with AMD throughout the entire protein structure. Overall, given the availability of a high resolution crystal structure for CFI, and a high proportion of rare coding variants affecting FI secretion, we conclude that our analysis shows high predictive value for interpreting CFI rare coding variants.

C9 polymerization plays a role in formation of the membrane attack complex, which, upon successful assembly, can disrupt the membrane of a target cell. Rare coding variants in C9 affect the polymerization of C9^14,18^ Here we observe POKEMON clusters in the LDLRA and MACPF domain of C9, further, of 25 tested variants 10 show a ddG above 2. For four of our testewd variants functional testing has been reported previously, the variant p.Thr170Ile is predicted to be highly disruptive with a high ddG of 45.95, in line with this significantly reduced secretion levels have been reported *in vitro* for this variant^14^. The variants p.Gly126Arg and p.Ala529Thr did not affect C9 secretion levels, while the ddg score predicts them to be disruptive and benign respectively. The variant p.Phe62Ser is presented to affect stability with a ddg of 6.12, however this variant showed significantly increased secretion *in vitro* ^14^.

Overall performing functional analysis of rare coding variants is time consuming and expensive, consequently accurate prediction models are valuable tools in the diagnostic setting. This process of identifying areas where protein stability plays a large role, depending on the DNA sequence that creates the protein, will be invaluable in finding biomarkers or targets for new treatment of disease. Clinical trials will find it useful to investigate targeted inclusion of rare variant carriers for complement inhibitors or other disease/protein trials, and will help with understanding of functional consequences of real world translational work.

## Data Availability

The genotype data analyzed during the current study were generated by the IAMDGC and are available through the database of Genotypes and Phenotypes (dbGAP; Accession: phs001039.v1.p1). Summary statistics for the IAMDGC data is available currently at http://amdgenetics.org.

## Author Contributions

MG, dJS, ELP, BJ, DR, CM, and JAC analyzed data. MG, dJS, ELP drafted and critically reviewed the manuscript. WSB, JLH, AIdH developed the ideas, advised on results and critically reviewed the manuscript.

## Ethical approval

The IRB of Case Western Reserve University – University Hospitals (IRB Number EM-14-04) gave ethical approval for this work.

The study participants were previously ascertained by IAMDGC cohorts as described in Fritsche et al, 2016, Nature Genetics. All participants provided informed consent, and the study was approved by institutional review boards as previously described

## Supplementary Figures and Tables

**Supplementary Figures 1A-D:**
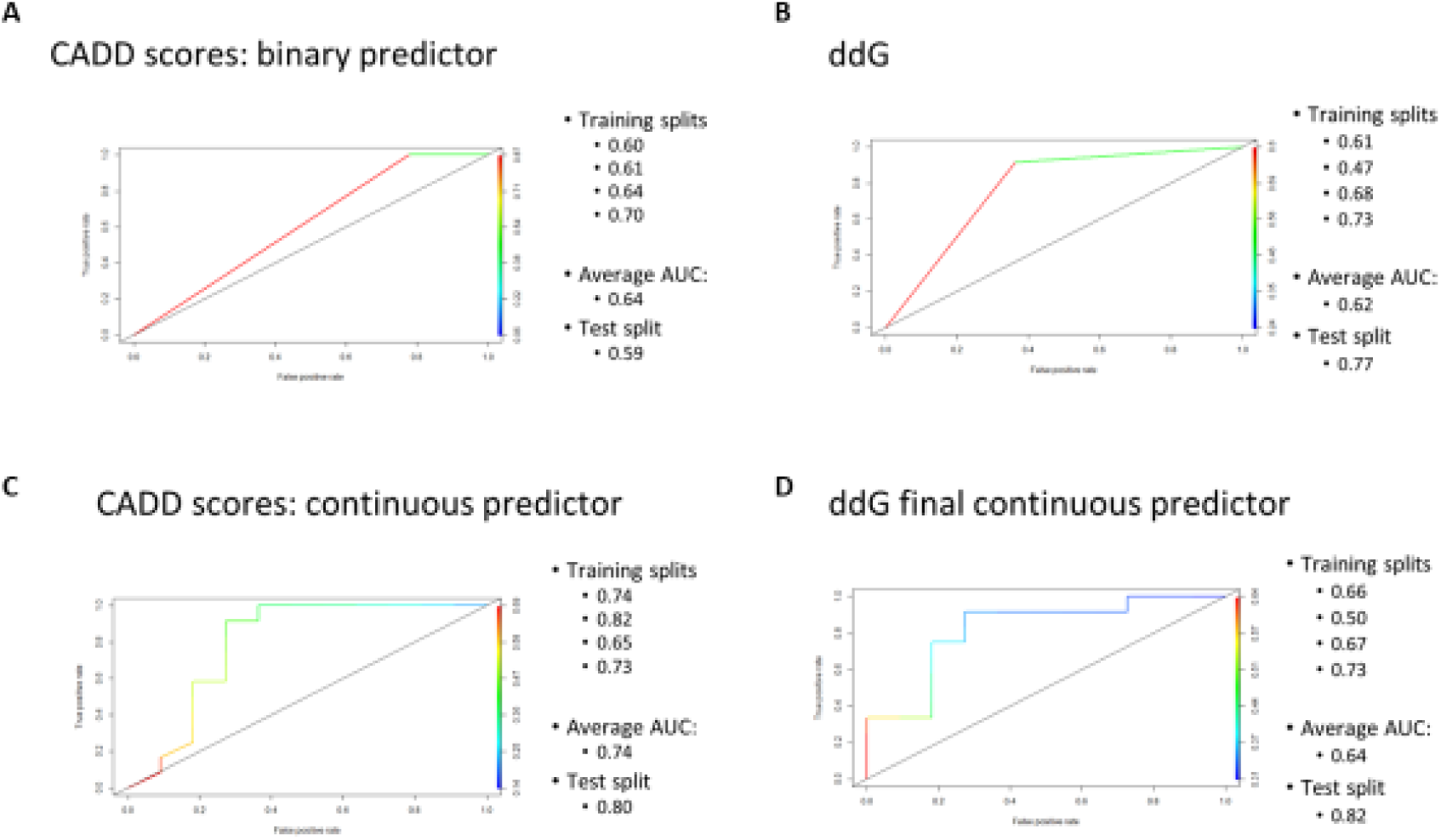
CADD and ddg scores both binary and continuous visuals.

Supplementary Table 1:

**Supplementary Table 1:** XX
|  |
| --- |
| **Variant with Plasma Available: Gene/Protein Variant** |
| CFI P273R |
| CFI T84A |
| CFI I518T |
| C3 I815M |
| C3 S219C |
| C3 T220I |
| CFB G252S |
| CFB V669L |
| CFB I569L |
| C9 I203V |
| C9 E300K |
| C9 F338L |
| C9 V467F |
| C9 L468F |
| C9 I476V |

Supplementary Table 2:

**Supplementary Table 2:**
| Target | FI | FB | C3bBbP | sTCC |
| --- | --- | --- | --- | --- |
| Coating | 1000x diluted sheep anti-human FI antibody (LabNed, Netherlands) | 2000x diluted polyclonal anti-human FB antibody (antibodies-online.com, Germany) | 1000x diluted mouse anti-human Factor P#2 (Quidel, USA) | 200x diluted anti-C5b-9 antibody (bioporto diagnostics, Denmark) |
| Blocking* | First with SuperBlock (ThermoFisher Scientific, USA), and then with 1% bovine serum albumin (BSA, Sigma, USA) | 1% BSA (Sigma, USA) | 1% BSA (Sigma, USA) | 1% BSA (Sigma, USA) |
| Samples | 1000x, 2000x and 4000x diluted in PBST 0.2% BSA | 4000x and 8000x diluted in PBST 0.2% BSA | 50x in PBST-10mM EDTA | 200x in PBST-10mM EDTA |
| Detection antibody | 1000x mouse anti-human FI (ProSci, USA) | 8000x diluted anti-human FB HRP-conjugated (antibodies-online.com, Germany) | 2000x diluted anti-human C3c (Siemens, Germany) | 2000x diluted anti-human C6, biotinylated (Quidel, USA) |
| Secondary antibody | goat anti-mouse HRP (DAKO, Germany) | na | Goat anti-rabbit HRP (DAKO, Germany) | na |
| Development | o-phenylenediamine dihydrochloride (OPD) substrate (Dako, Germany) | Tetra Methyl Benzidine (TMB) substrate (Sigma, USA) | OPD substrate (Dako, Germany) | Streptavidin-HRP (Pierce Biotechnology, USA) |
| * in between each step after blocking, the plates were washed four times with 200µl PBS 0.02% Tween |  |  |  |  |

**Supplementary Table 3:**
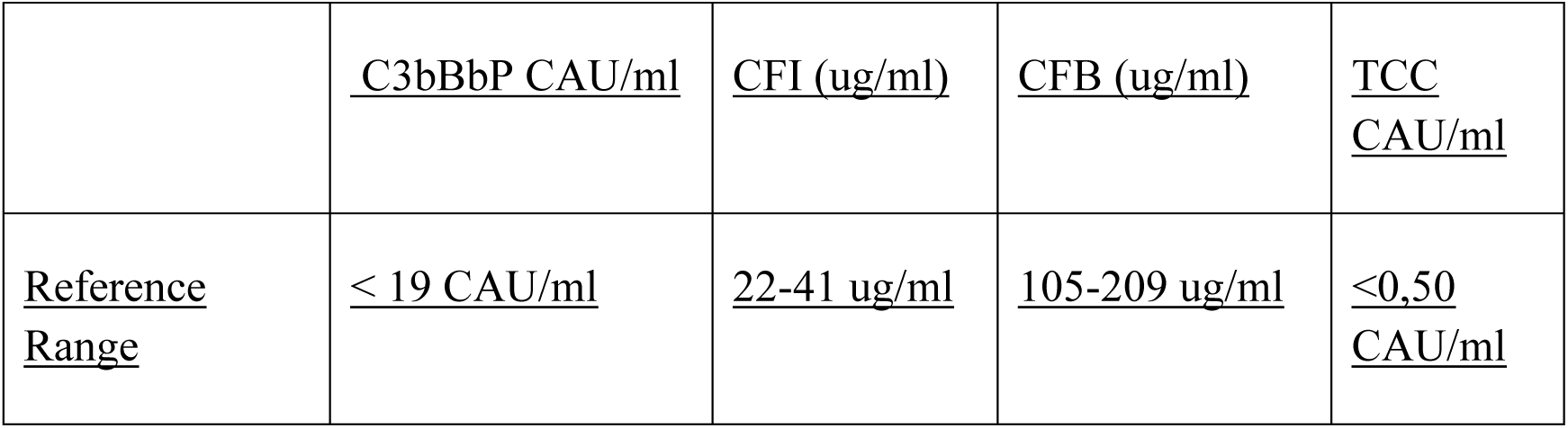
ELISA reference values.

|  |  |  |  |  |
| --- | --- | --- | --- | --- |
|  | <u>C3bBbP CAU/ml</u> | <u>CFI (ug/ml)</u> | <u>CFB (ug/ml)</u> | <u>TCC CAU/ml</u> |
| <u>Reference Range</u> | <u>< 19 CAU/ml</u> | <u>22-41 ug/ml</u> | <u>105-209 ug/ml</u> | <u><0,50 CAU/ml</u> |

**Supplementary Table 4:**
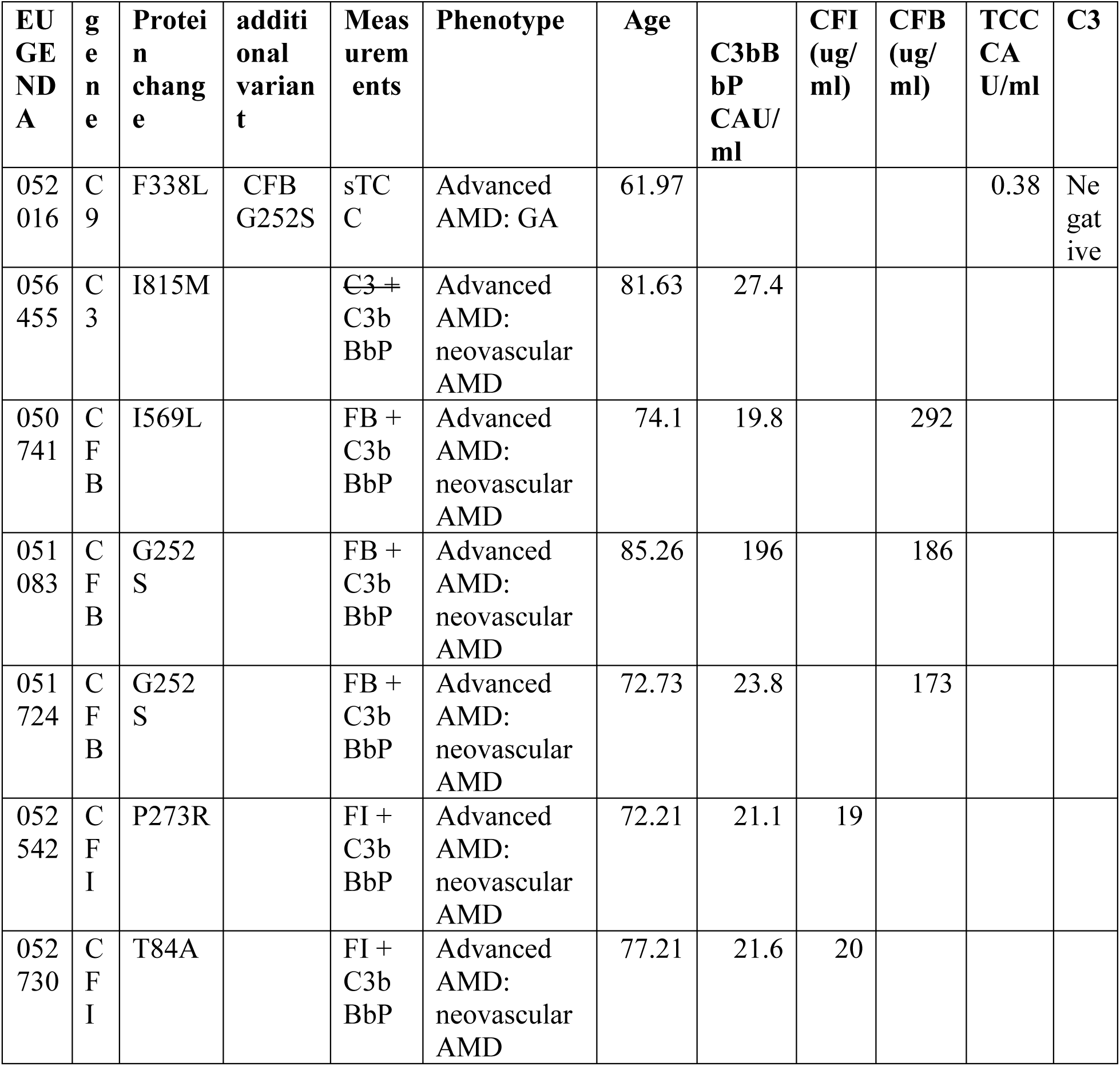
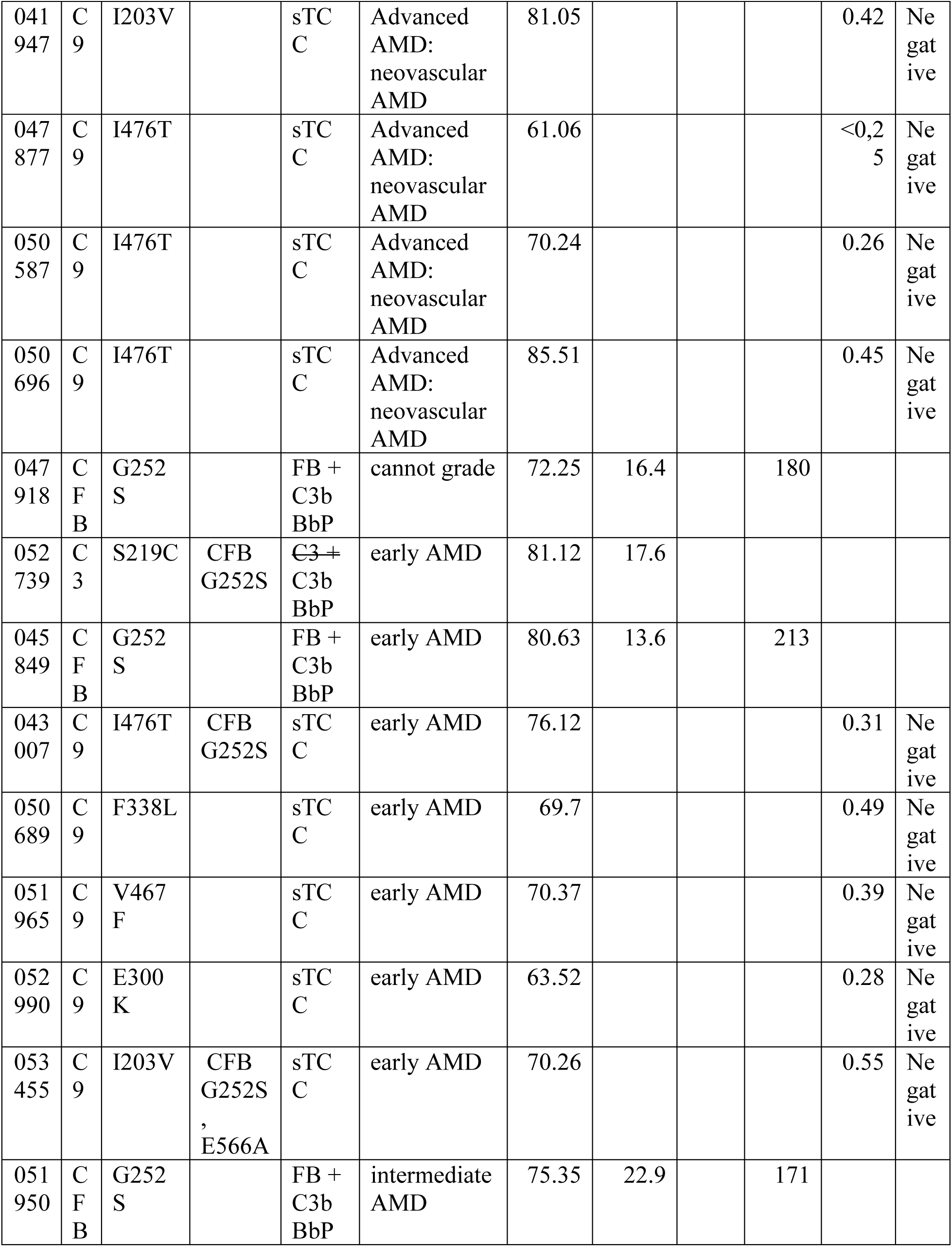
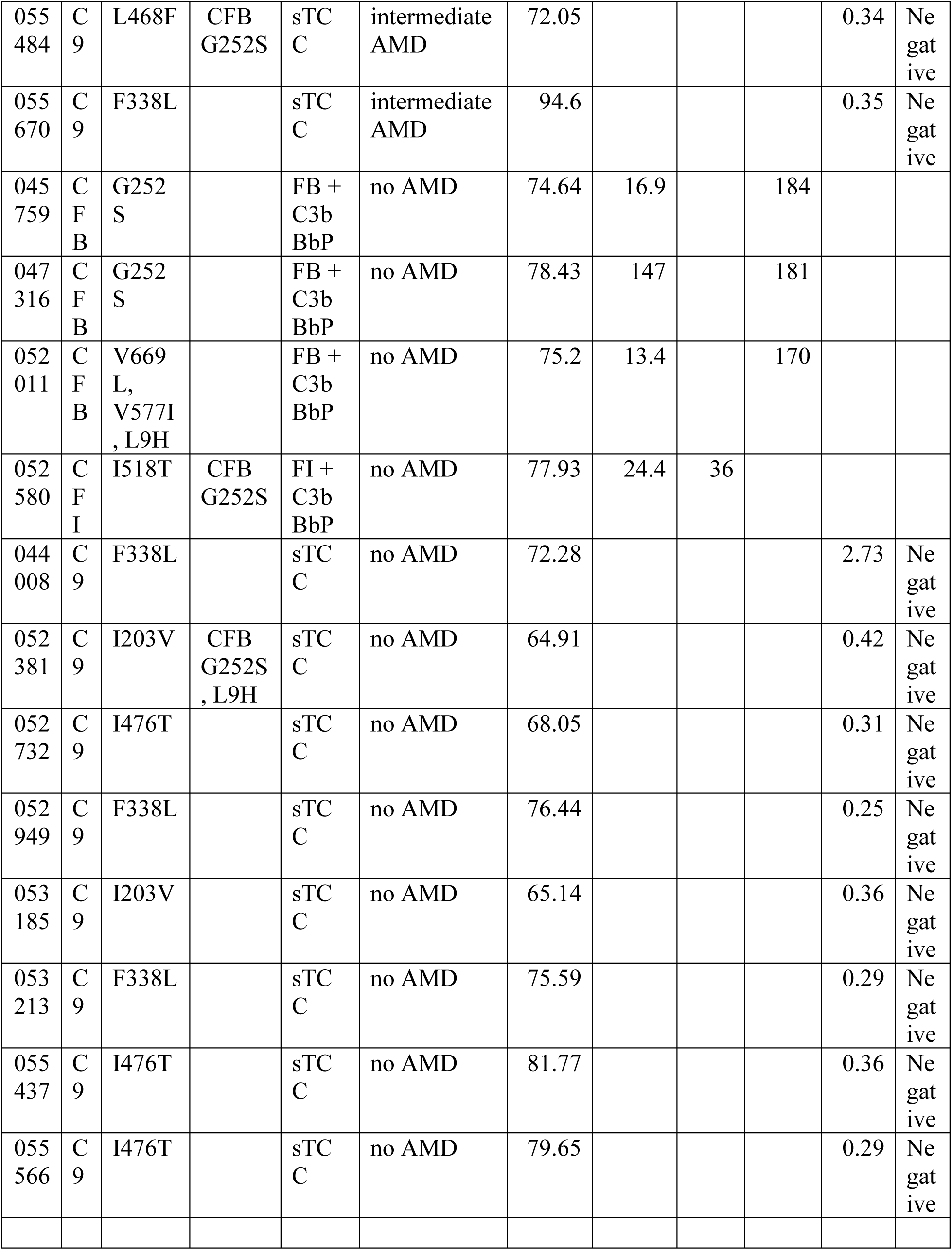
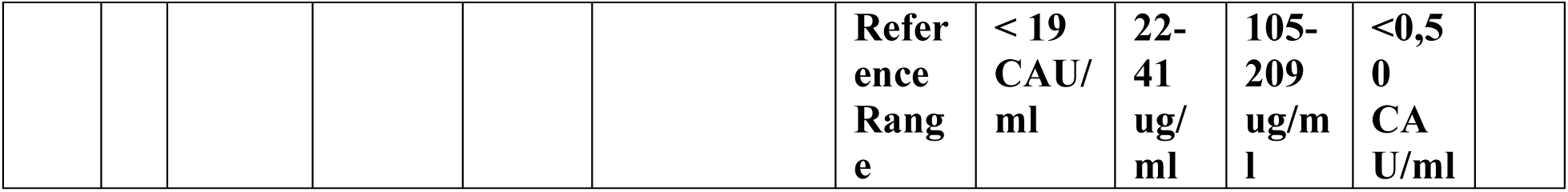
All patient ID, genes tested, and values of ELISA on complement gene pathway outcomes.

## References

1. Grunin, M. et al. Integrating Computational Approaches to Predict the Effect of Genetic Variants on Protein Stability in Retinal Degenerative Disease. Adv. Exp. Med. Biol. 1415, 157–163 (2023).

2. Flanagan, S. E., Patch, A.-M. & Ellard, S. Using SIFT and PolyPhen to predict loss-of-function and gain-of-function mutations. Genet. Test. Mol. Biomarkers 14, 533–537 (2010).

3. Kucukkal, T. G. & Alexov, E. Structural, dynamical, and energetical consequences of RETT syndrome mutation R133c in MeCP2. Comput. Math. Methods Med. 2015, (2015).

4. Petukh, M., Kucukkal, T. G. & Alexov, E. On human disease-causing amino acid variants: Statistical study of sequence and structural patterns. Hum. Mutat. 36, (2015).

5. Yue, P., Li, Z. & Moult, J. Loss of protein structure stability as a major causative factor in monogenic disease. J. Mol. Biol. 353, (2005).

6. Christ, C. D., Mark, A. E. & van Gunsteren, W. F. Basic ingredients of free energy calculations: A review. J. Comput. Chem. 31, 1569–1582 (2010).

7. Kumar, M. D. S. et al. ProTherm and ProNIT: thermodynamic databases for proteins and protein-nucleic acid interactions. Nucleic Acids Res. 34, D204–6 (2006).

8. Burley, S. K. et al. Protein Data Bank (PDB): The Single Global Macromolecular Structure Archive. Methods Mol. Biol. 1607, 627–641 (2017).

9. Waterhouse, A. et al. SWISS-MODEL: Homology modelling of protein structures and complexes. Nucleic Acids Res. 46, (2018).

10. Fritsche, L. G. et al. A large genome-wide association study of age-related macular degeneration highlights contributions of rare and common variants. Nat. Genet. 48, 134– 143 (2016).

11. van de Ven, J. P. H. et al. A functional variant in the CFI gene confers a high risk of age-related macular degeneration. Nat. Genet. 45, 813–817 (2013).

12. Kavanagh, D. et al. Rare genetic variants in the CFI gene are associated with advanced age-related macular degeneration and commonly result in reduced serum factor I levels. Hum. Mol. Genet. 24, 3861–3870 (2015).

13. Geerlings, M. J., Kersten, E., Groenewoud, J. M. M., Fritsche, L. G. & Hoyng, C. B. Geographic distribution of rare variants associated with age-related macular degeneration. Mol. Vis. 9, 75–82 (2018).

14. Kremlitzka, M. et al. Functional analyses of rare genetic variants in complement component C9 identified in patients with age-related macular degeneration. Hum. Mol. Genet. 27, (2018).

15. de Jong, S. et al. Effect of rare coding variants in the CFI gene on Factor I expression levels. Hum. Mol. Genet. 29, (2020).

16. Java, A. et al. Functional Analysis of Rare Genetic Variants in Complement Factor I (CFI) using a Serum-Based Assay in Advanced Age-related Macular Degeneration. Transl. Vis. Sci. Technol. 9, 37 (2020).

17. Hallam, T. M. et al. Rare genetic variants in complement factor i lead to low FI plasma levels resulting in increased risk of age-related macular degeneration. Investig. Ophthalmol. Vis. Sci. 61, (2020).

18. McMahon, O. et al. The rare C9 P167S risk variant for age-related macular degeneration increases polymerization of the terminal component of the complement cascade. Hum. Mol. Genet. 30, 1188–1199 (2021).

19. Wong, E. K. S. et al. Functional Characterization of Rare Genetic Variants in the N-Terminus of Complement Factor H in aHUS, C3G, and AMD. Front. Immunol. 11, 602284 (2020).

20. de Jong, S. et al. Systemic complement levels in patients with age-related macular degeneration carrying rare or low-frequency variants in the CFH gene. Hum. Mol. Genet. 31, 455–470 (2022).

21. Sivley, R. M., Dou, X., Meiler, J., Bush, W. S. & Capra, J. A. Comprehensive Analysis of Constraint on the Spatial Distribution of Missense Variants in Human Protein Structures. Am. J. Hum. Genet. 102, (2018).

22. Sivley, R. M. et al. Three-dimensional spatial analysis of missense variants in RTEL1 identifies pathogenic variants in patients with Familial Interstitial Pneumonia. BMC Bioinformatics 19, (2018).

23. Jin, B., et al. An Association Test of the Spatial Distribution of Rare Missense Variants within Protein Structures Improves Statistical Power of Sequencing Studies. bioRxiv (2021).

24. McCarthy, S. et al. A reference panel of 64,976 haplotypes for genotype imputation. Nat. Genet. (2016) doi:10.1038/ng.3643.

25. Delaneau, O., Marchini, J. & Zagury, J.-F. A linear complexity phasing method for thousands of genomes. Nat. Methods 9, 179–181 (2012).

26. van Leeuwen, E. M. et al. Population-specific genotype imputations using minimac or IMPUTE2. Nat. Protoc. 10, 1285–1296 (2015).

27. Gao, F., et al. XWAS: a software toolset for genetic data analysis and association studies of the X chromosome. bioRxiv 009795 (2014) doi:10.1101/009795.

28. Karczewski, K. J. et al. The mutational constraint spectrum quantified from variation in 141,456 humans. Nature 581, (2020).

29. Karczewski, K. J. et al. The ExAC browser: displaying reference data information from over 60 000 exomes. Nucleic Acids Res. 45, D840–D845 (2017).

30. Pearson, W. R. Selecting the Right Similarity-Scoring Matrix. Curr. Protoc. Bioinforma. 43, 3.5.1–3.5.9 (2013).

31. Alford, R. F. et al. The Rosetta All-Atom Energy Function for Macromolecular Modeling and Design. J. Chem. Theory Comput. 13, (2017).

32. Kellogg, E. H., Leaver-Fay, A. & Baker, D. Role of conformational sampling in computing mutation-induced changes in protein structure and stability. Proteins 79, 830– 838 (2011).

33. Chen, H. et al. Sequence kernel association test for survival traits. Genet. Epidemiol. 38, 191–197 (2014).

34. Jin, B. et al. An association test of the spatial distribution of rare missense variants within protein structures identifies Alzheimer’s disease-related patterns. Genome Res. 32, 778– 790 (2022).

35. de Jong, S. et al. Functional Analysis of Variants in Complement Factor I Identified in Age-Related Macular Degeneration and Atypical Hemolytic Uremic Syndrome. Front. Immunol. 12, 789897 (2021).

